# Cluster-Based Toxicity Estimation of Osteoradionecrosis via Unsupervised Machine Learning: Moving Beyond Single Dose-Parameter Normal Tissue Complication Probability by Using Whole Dose-Volume Histograms for Cohort Risk Stratification

**DOI:** 10.1101/2023.03.24.23287710

**Authors:** Seyedmohammadhossein Hosseinian, Mehdi Hemmati, Cem Dede, Travis C. Salzillo, Lisanne V. van Dijk, Abdallah S. R. Mohamed, Stephen Y. Lai, Andrew J. Schaefer, Clifton D. Fuller

## Abstract

**Purpose:** Given the limitations of extant models for normal tissue complication probability estimation for osteoradionecrosis (ORN) of the mandible, the purpose of this study was to enrich statistical inference by exploiting structural properties of data and provide a clinically reliable model for ORN risk evaluation through an unsupervised-learning analysis.

**Materials and Methods:** The analysis was conducted on retrospective data of 1,259 head and neck cancer (HNC) patients treated at the University of Texas MD Anderson Cancer Center between 2005 and 2015. The (structural) clusters of mandibular dose-volume histograms (DVHs) were identified through the K-means clustering method. A soft-margin support vector machine (SVM) was used to determine the cluster borders and partition the dose-volume space. The risk of ORN for each dose-volume region was calculated based on the clinical risk factors and incidence rates.

**Results:** The K-means clustering method identified six clusters among the DVHs. Based on the first five clusters, the dose-volume space was partitioned almost perfectly by the soft-margin SVM into distinct regions with different risk indices. The sixth cluster overlapped the others entirely; the region of this cluster was determined by its envelops. These regions and the associated risk indices provide a range of constraints for dose optimization under different risk levels.

**Conclusion:** This study presents an unsupervised-learning analysis of a large-scale data set to evaluate the risk of mandibular ORN among HNC patients. The results provide a visual risk-assessment tool (based on the whole DVH) and a spectrum of dose constraints for radiation planning.

## 1. Introduction

Osteoradionecrosis (ORN) of the mandible is a debilitating side effect of radiation therapy for head and neck cancer (HNC) patients [1, 2, 3, 4, 5]. ORN is commonly defined as non-healing bone for a period of at least three months due to exposure to radiation [6, 7, 8]. Despite its relatively low prevalence [9, 10, 11], ORN may severely affect the quality of life of surviving HNC patients; it is often accompanied by pain, dysaesthesia, dysgeusia, and difficulties in mastication and speech, and in case of progression, it may lead to infection, bone fracture, and intraoral or extra-oral fistulae [1, 6, 12, 13, 14]. With the improved life expectancy of HNC patients, especially among young adults with human papillomavirus associated tumors [11], the importance of ORN prevention strategies in modern practice is spotlighted more than ever before. This requires developing reliable risk evaluation models for ORN to guide dose optimization/adaptation in radiation therapy planning.

Normal tissue complication probability (NTCP) is a widely accepted prediction tool for radiation toxicity [15, 16, 17, 18, 19]. NTCP models aim to provide the likelihood of treatment-induced complications based on the radiation dose delivered to organs at risk (OARs) as well as other clinical risk factors. The current paradigm of NTCP modeling for HNCs relies on supervised-learning (classification) methods, most popularly the classical *logistic regression* method [20].

During the past decade, a vast number of classification-based NTCP models have been proposed for various HNC treatment-induced complications [11, 21, 22, 23, 24, 25, 26, 27, 28, 29]. The classification methods, however, have a major limitation for NTCP modeling—due to *multicollinearity* of dose features—which undermines clinical reliability of these models.

Distribution of radiation dose over an OAR is commonly represented by a dose-volume histogram (DVH). As an OAR’s DVH depicts a *distribution*, the dose-related features for these models (i.e., DVH values) are highly correlated. As a result, the entire DVH curve is summarized into a single number (e.g., mean dose), which completely disregards the *distribution* of radiation dose and the fact that clinically different DVH curves—with different toxicity outcomes—may share a feature value. Single-parameter evaluation of dose distributions has been reported as a caveat of early NTCP models (e.g., LKB) for clinical use [16], yet it has persisted throughout the subsequent data-driven models, due to the multicollinearity issue.

In addition, the best performance of classification methods concerns *separable* data [30]. In general, these methods aim to learn the best separator of two training classes (i.e., safe radiation plans vs. those leading to complications) from observed clinical data. They perform well even if the data is softly separable (i.e., with a relatively few violations), but in case of inseparable (i.e., highly mixed) classes, they generally do not perform better than a *naive Bayes* classifier. Naive Bayes is a simple classifier whose predictions mainly rely on the observed incidence rates [30]. The effect of data inseparability on classification accuracy is shown in Appendix A. Based on available data of mandibular ORN development among HNC patients [31], there exists evidence that the two classes of ORN and non-ORN patients are highly inseparable by radiation dose to the mandible; see Appendix B. The inseparability observation is aligned with an evidence-based pathogenesis theory of ORN, suggesting that radiation arteritis leads to the development of a hypocellular, hypovascular, and hypoxic environment, and consequently ORN [6, 7]. This implies that a decisive factor in ORN development is the radiation dose received by mandibular arteries, including arterioles and capillaries. Capturing the dose delivered to such a fine network is impractical with the current technologies. On the other hand, DVHs summarize the spatially (3D) distributed radiation dose into one-dimensional data. For statistical inference, such a “summary” is a sufficient statistic only if radiosensitivity of the OAR is spatially homogeneous. However, homogeneous distribution of arteries across the mandible is an unrealistic assumption. This suggests that, while mandibular DVHs are the best available clinical indicator of ORN development, they generally do not contain sufficient information to separate the two classes (ORN vs. non-ORN) rigorously as intended in the advanced classification methods.

The aforementioned limitations of supervised-learning methods motivate alternative analyses to build more informative and clinically reliable models for evaluation of the risk of ORN. This paper presents an unsupervised-learning approach for ORN risk assessment and the results of the analysis on retrospective data of a large cohort of HNC patients.

## 2. Materials and Methods

### 2.1 Data

The analysis was conducted on a data set compiled and made publicly available by van Dijk et al. [31]. The data is composed of anonymized (retrospective) clinical records and the mandibular DVHs of 1,259 HNC patients treated by radiation therapy alone or in combination with surgery and/or chemotherapy at the University of Texas MD Anderson Cancer Center between 2005 and 2015, following an institutional review board approval (RCR030800). The clinical records include age, gender, cancer subsite, T-stage, N-stage, pre-radiation (within 6 weeks before treatment) dental extraction status, smoking status and pack-years, chemotherapy treatment, definitive vs. postoperative radiation therapy, and the mandible bone volume. The reported DVH values include D2%, D5% to D95% in 5% increments, D97%, D98%, and D99% as well as V5Gy to V70Gy in 5 Gy increments; the minimum, maximum, and mean dose (Gy) delivered to the mandible are also included. Out of the 1,259 patients, 1,086 (86.3%) have been reported ORN-free during the minimum of 12 months of post-therapy follow-up; 173 (13.7%) patients are reported to have developed ORN (grade I to IV) during the follow-up period. A complete description of the data, including more information about the cohort and treatments as well as the details of data extraction and processing, has been presented in [11]. The data is publicly available at https://doi.org/10.6084/m9.figshare.13568207.

### 2.2 Analysis

This paper presents a secondary analysis of the data set described above; van Dijk et al. [11] have used this data to train a multivariate logistic regression NTCP model for mandibular ORN (any grade). Their univariate analysis, followed by AIC-based forward stepwise selection, has identified D30% and pre-radiation dental extraction (PDE) status as the model features; PDE is a nominal (binary) feature indicating no/edentulous vs. dental extractions. While their statistical analysis rigorously identifies radiation intensity and dental extraction as significant predictive factors of ORN development, due to multicollinearity of the dose features, the DVH curve is reduced to D30% in their model. Besides, our analysis shows that the performance of the proposed logistic regression model is almost equivalent to that of naive Bayes (AUC of 0.77 for logistic regression vs. 0.75 for naive Bayes); see Appendix C. This confirms that, due to inseparability of classes, the logistic regression predictions are mainly driven by the ORN incidence rates with respect to the model features. Based on this observation, we present a cluster-based analysis that provides more clinical insight about the risk of ORN development as a function of radiation intensity and dental extraction status.

Our method takes advantage of the structural properties of mandibular DVHs to divide them into disjoint subgroups (of similar radiation plans), each spanning a distinct region of the dose-volume space. It then renders a risk index for each region based on the observed incidence rate of ORN (depending on PDE status) in the associated DVH subgroup. The analysis was conducted in three steps, described below. Steps 1 and 2 were performed using the scikit-learn library of Python [32].

**Step 1:** The K-means clustering method [30] was employed to identify the inherent clusters of the mandibular DVHs. To this end, each DVH curve was represented as an array of dose values (i.e., Dx% for x% ranging from 2% to 99%); the inertia (within-cluster sum-of-squares) criterion was used to identify an optimal number of clusters.

**Step 2:** To partition the dose-volume space with respect to these clusters, the shared borders of adjacent clusters were identified by a soft-margin support vector machine (SVM) [30]. The analysis was conducted pointwise (for each Dx% individually) to obtain smooth borders among the clusters with minimal violations.

**Step 3:** For each region of the dose-volume space, spanned by an identified cluster, the ORN incidence rate (within the cluster) per PDE status was reported as the associated risk index. As a result, each dose-volume region was associated with two risk indices, for no/edentulous and dental extractions, separately.

## 3. Results

The K-means clustering method identified 5-6 clusters among the mandibular DVHs. In general, as the number of clusters (K) increases, the clustering inertia decreases; an optimal number of clusters is identified by the value of K after which the inertia curve behaves linearly. Figure 1 illustrates the inertia plot of our analysis. Based on this figure, K = 5 is optimal; however, the sixth cluster identified by the clustering method (through K = 6) is structurally different from the first five clusters and provides additional insight. Therefore, we considered both cases.

**Figure 1:**
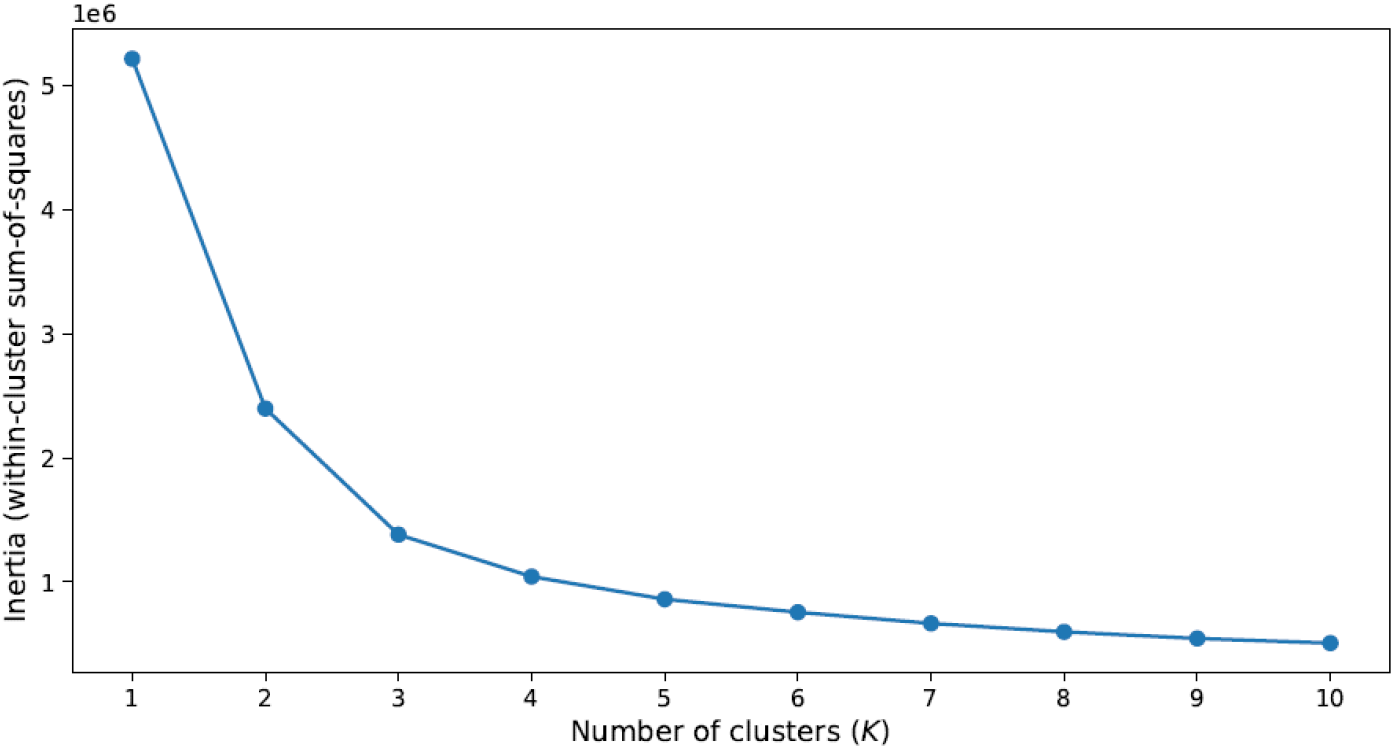
Inertia plot of the K-means clustering method

Figure 2 illustrates the identified clusters by the K-means clustering method, color-coded for better visualization. We have also represented DVHs directly by Dx% values (in Gy) rather than the conventional dose-volume axes, to increase readability. An important distinction between the results for K = 5 and K = 6 concerns the sixth cluster, depicted in black in Figure 2b. This cluster is mainly composed of a group of DVHs with high dose (above 60 Gy) almost constantly imposed over most of the mandible volume. While this group is visually distinguishable from the other clusters, with K = 5, it has been identified as a part of the top cluster, illustrated in red, implying it is most similar to that cluster structurally.

**Figure 2:**
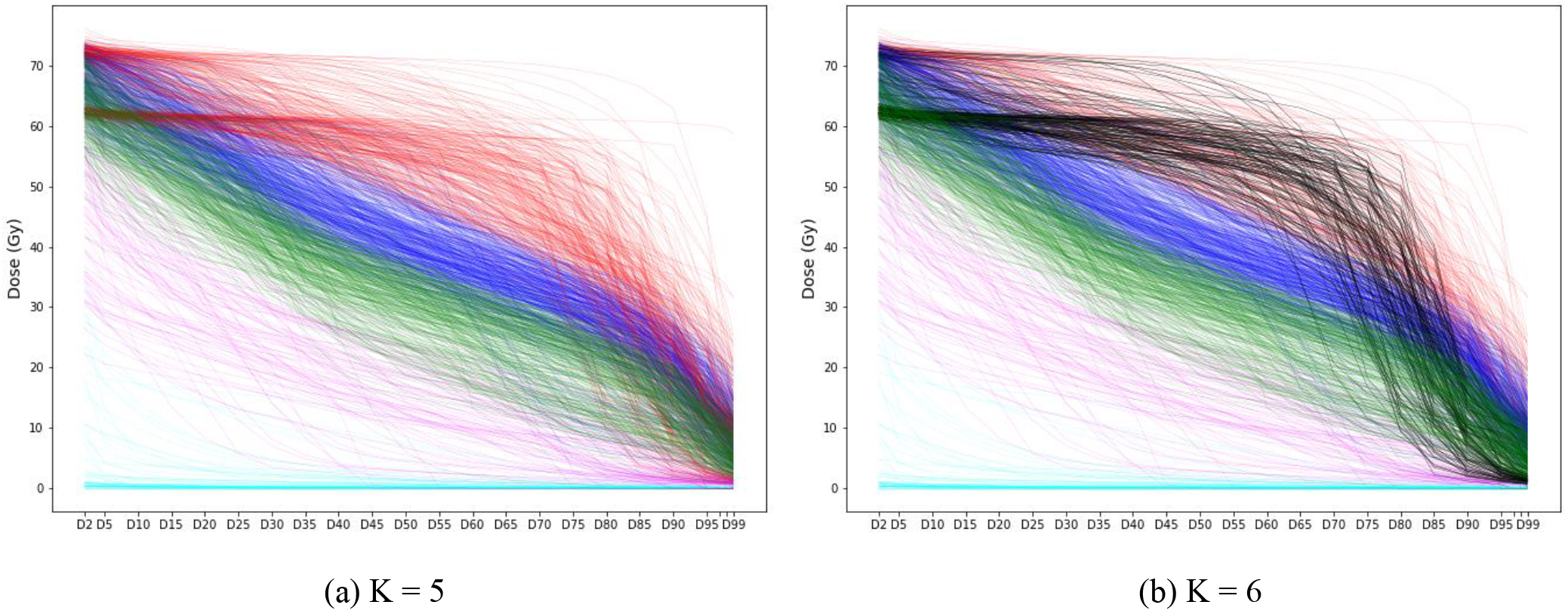
Visualization of the identified clusters

As the next step of the analysis, a soft-margin SVM was employed to identify the cluster borders and partition the dose-volume space; SVM renders the best separator of two classes. As the analysis was performed pointwise, the best separating point (with minimum violations) was identified for every pair of adjacent clusters for each Dx%. With K = 5, as the black cluster was considered a part of the red cluster and highly overlapping with the others in the most left and right parts of the plot, SVM failed to identify its border in the [D2%, D5%] and [D90%, D99%] intervals. However, with K = 6 and exclusion of the sixth (black) cluster, SVM was able to identify the cluster borders and partition the dose-volume space almost perfectly. For the sixth cluster, we determined the corresponding dose-volume region by its envelopes (i.e., pointwise minimum and maximum dose). Figure 3 illustrates the cluster borders by dashed black lines. As the choice of K = 6 provided a more accurate partition of the dose-volume space, we present the final step of the analysis with six clusters.

**Figure 3:**
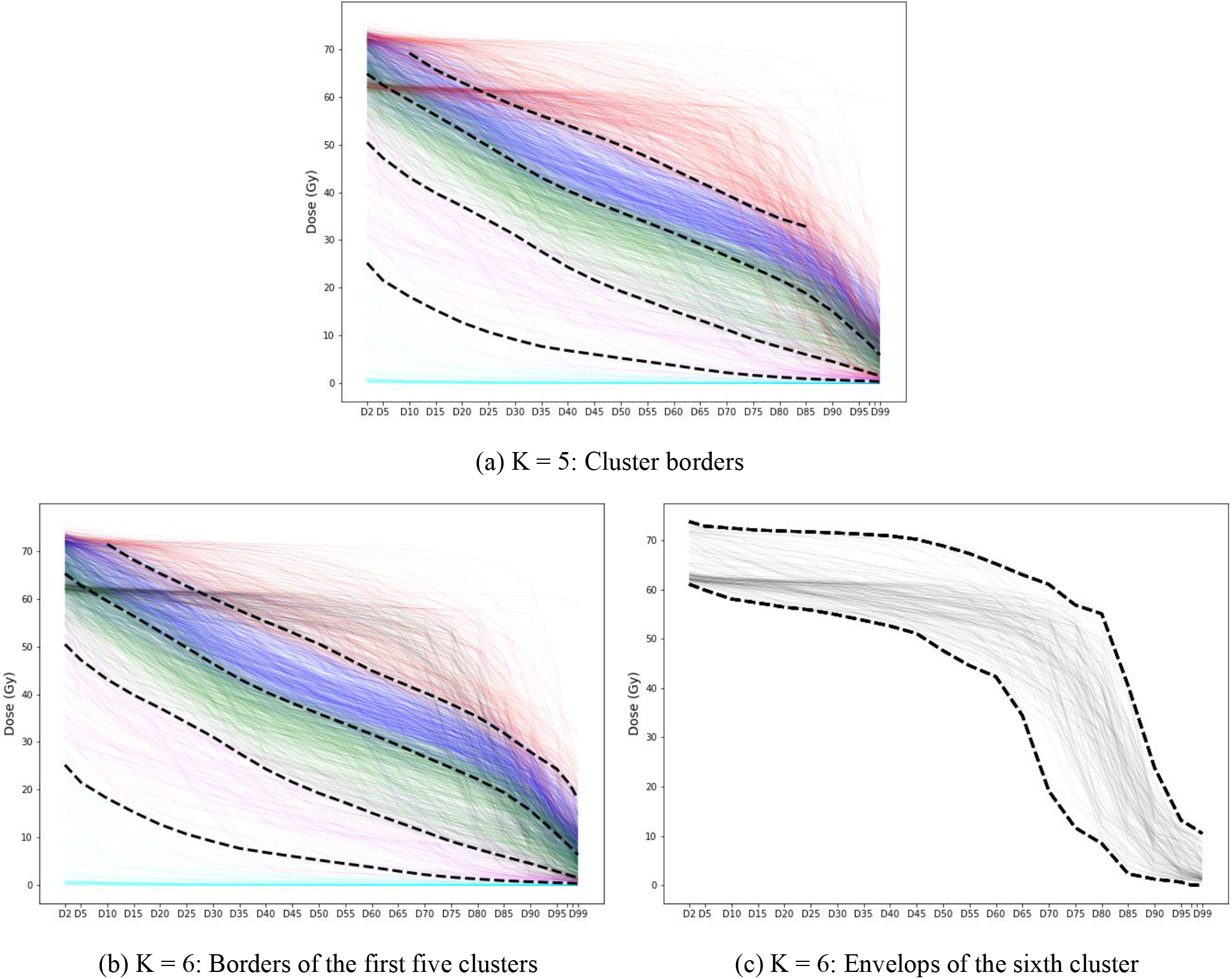
Cluster borders identified by a soft-margin SVM

For each part of the dose-volume space, characterized by the borderlines of the identified clusters, two risk indices were calculated based on the observed incidence rate of ORN in the corresponding cluster per PDE status. Table 1 reports the observed frequency of ORN development among the patients for each cluster per PDE status. In this table, PDE = 0 indicates no/edentulous dental extractions, and PDE = 1 concerns performed dental extractions. We have labeled the first five clusters according to their order from the lower left to the upper right corners of the DVH plot in Figure 3b; cluster No. 6 is the overlapping cluster, illustrated in black in Figures 3b and 3c. The columns “ORN incidence” indicate the number of patients with a positive ORN record out of the total number of patients in that cluster with the same PDE status, and “Risk index (*r*)” is the corresponding ratio. Figure 4 shows the risk indices on the dose-volume regions identified in the previous step; each region spans the area between two consecutive cluster borders, except for the region of cluster No. 6, which was determined by its envelops. The borders of the sixth cluster are depicted by dotted lines in Figure 4.

**Table 1:**
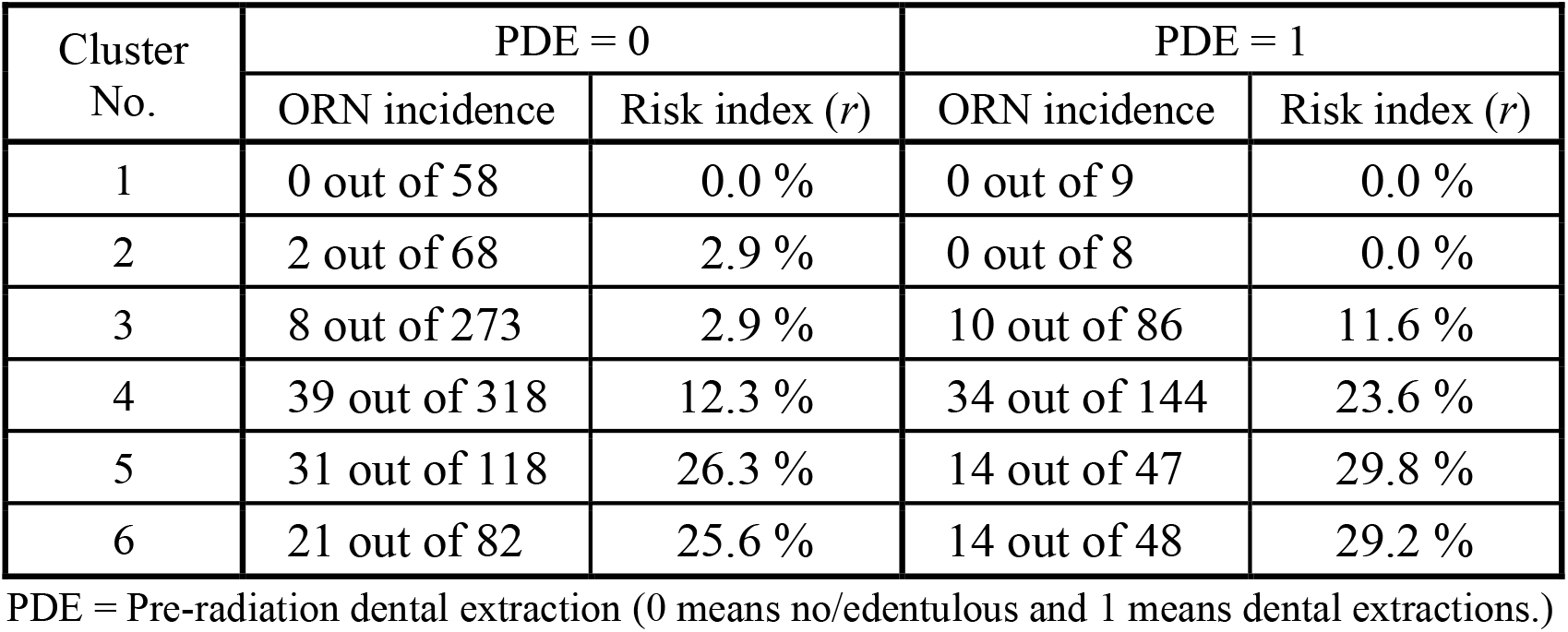
ORN incidence rates among the identified clusters per PDE status

| Cluster No. | PDE = 0 |  | PDE = 1 |  |
| --- | --- | --- | --- | --- |
| | ORN incidence | Risk index ($r$) | ORN incidence | Risk index ($r$) |
| 1 | 0 out of 58 | 0.0 % | 0 out of 9 | 0.0 % |
| 2 | 2 out of 68 | 2.9 % | 0 out of 8 | 0.0 % |
| 3 | 8 out of 273 | 2.9 % | 10 out of 86 | 11.6 % |
| 4 | 39 out of 318 | 12.3 % | 34 out of 144 | 23.6 % |
| 5 | 31 out of 118 | 26.3 % | 14 out of 47 | 29.8 % |
| 6 | 21 out of 82 | 25.6 % | 14 out of 48 | 29.2 % |

PDE = Pre-radiation dental extraction (0 means no/edentulous and 1 means dental extractions.)

**Figure 4:**
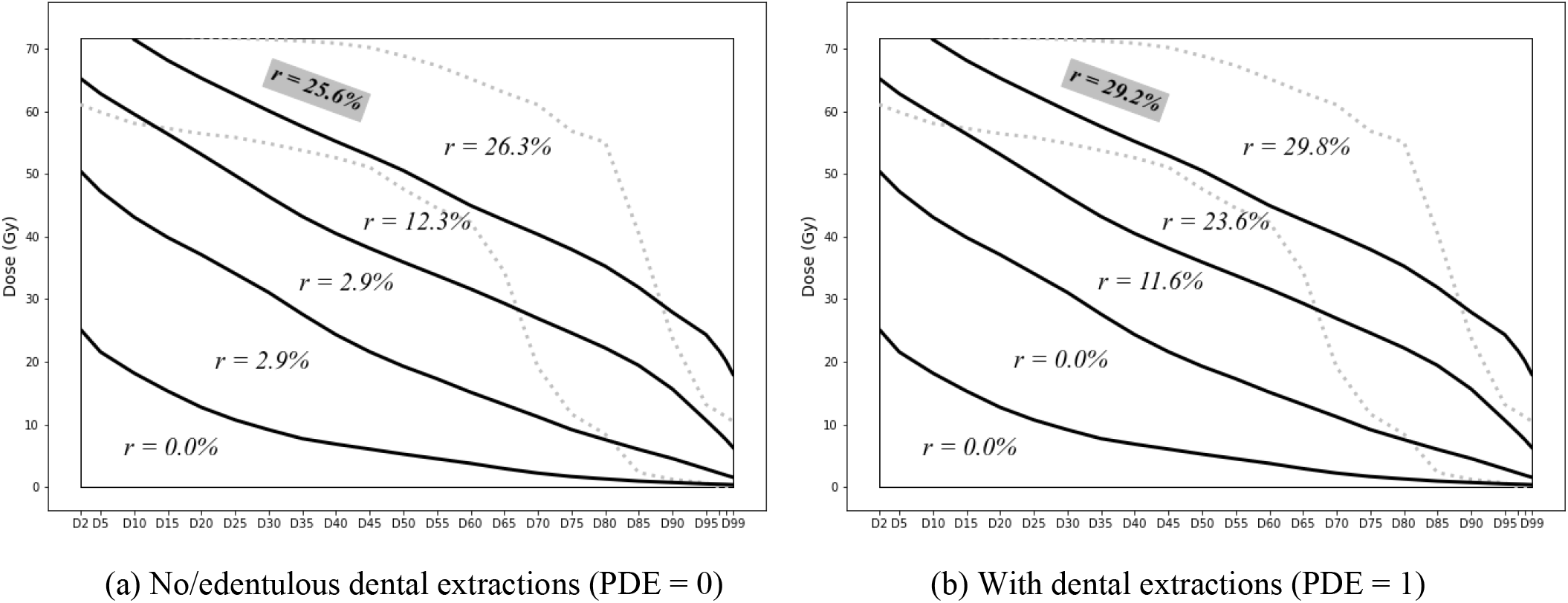
Risk indices of dose-volume regions

The results clearly show significant correlations between the risk of ORN and radiation intensity as well as PDE status, which is aligned with the univariate analysis results and confirms the relevance of structural clusters to the risk of ORN development. As mentioned earlier, cluster No. 6 (black) is structurally most similar to cluster No. 5 (red); it was identified as a part of the red cluster with K = 5. The results also show that these two clusters have very similar risk indices in both PDE categories (25.6% vs. 26.3% for PDE = 0 and 29.2% vs. 29.8% for PDE = 1). This further reveals the merit of the analysis results. Our method provides a visual tool for ORN risk evaluation of radiation plans based on the region that its mandibular DVH falls in and the patient’s PDE status. As the risk is characterized by the whole DVH (not a single-value summary), the results provide a spectrum of dose constraints (i.e., limit on various Dx% values for dose optimization) to ensure that the risk of ORN development stays below a certain level.

## 4. Discussion

The main advantage of the presented analysis is evaluation of radiation plans based on the whole DVH. As mentioned earlier, due to the multicollinearity issue, DVHs are reduced to a single-value summary in classification-based NTCP models, which disregards the fact that clinically different DVH curves—with different toxicity outcomes—may share a feature value. This can be observed by the toxicity outcome for the patients whose D30% is in the range of 55 Gy to 60 Gy, in our data set. According to van Dijk et al. [11] model, the risk of ORN for these patients (per PDE status) is relatively the same, as their mandibular DVHs have close D30% values. However, the actual treatment outcomes for these patients are different and arguably determined by the shape of DVHs. Figure 5 shows these patients’ DVHs; the incidence of ORN among the blue DVHs is 13.6% (3 out of 22) and among the black DVHs is 23.5% (12 out of 51). Other than this important difference, the van Dijk et al. [11] and our models predict similar risks for these patients, i.e., 13.0%-19.0% by van Dijk et al. [11] compared with 12.3%-25.6% (of the blue and black clusters) by our model for PDE = 0 and 22.4%-31.2% by van Dijk et al. [11] compared with 23.6%-29.2% (of the blue and black clusters) by our model for PDE = 1.

**Figure 5:**
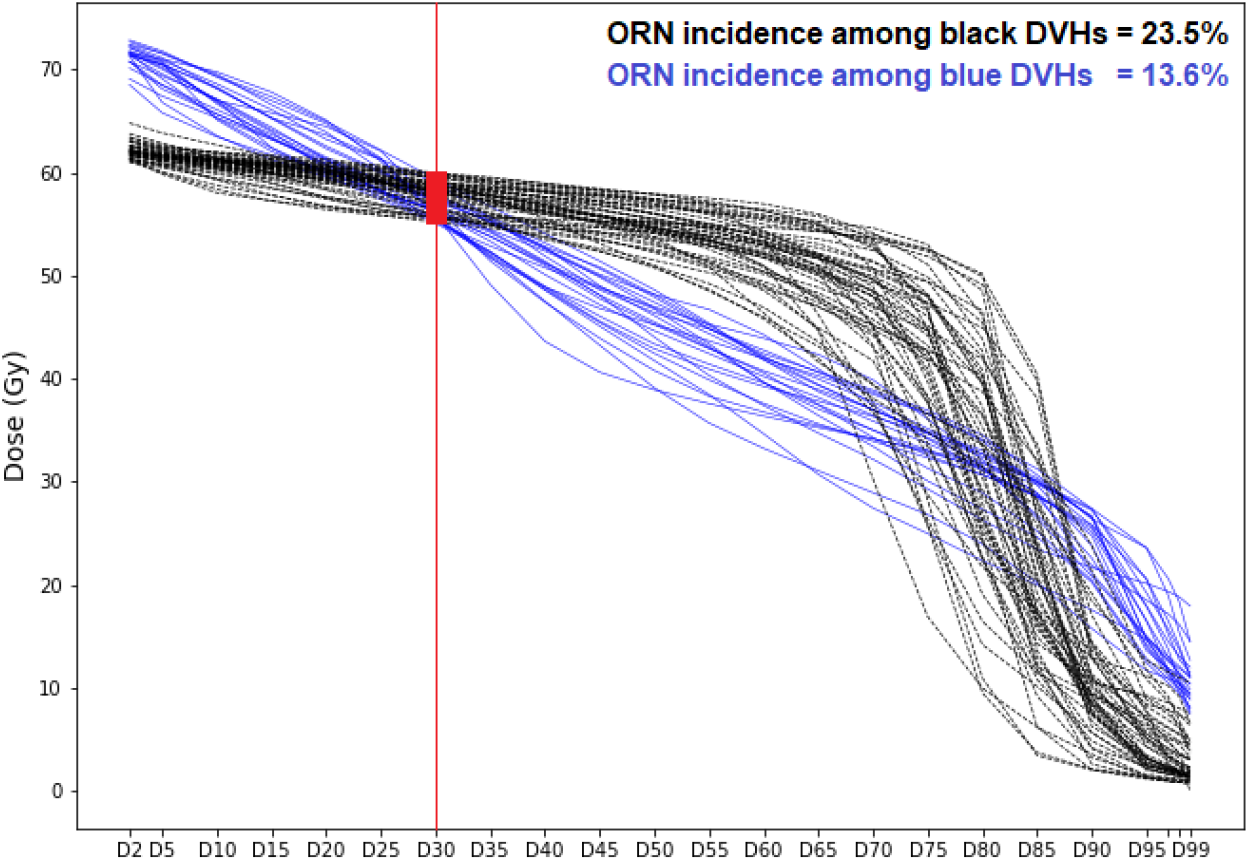
Different ORN incidences among DVHs with the same D30% value

Our motivation for this analysis method was the observation of inseparability of classes (i.e., ORN vs. non-ORN) by measurable features; see Appendix B. Incidence rate is the unique minimum-variance unbiased estimator (MVUE) for the mean of a Bernoulli random variable representing the hidden (unmeasurable) features. In the absence of information about the main separating features (i.e., highly inseparable classes by measurable features), such an estimator offers the most that can be learned from the data. This is supported by the close performances of logistic regression and naive Bayes classifiers on our data; see Appendix C. Those estimates will be naturally more accurate if the data is divided into smaller categories (clusters) characterized by statistically significant (measurable) features, to consider their correlations with the outcome. Thus, the presented analysis not only considers the whole DVH (not a single-value summary), but also enjoys a simple yet robust statistical-inference foundation.

The proposed approach has its own limitations to be considered. First, the choice of the number of clusters (K) in the K-means method is relatively subjective. While the inertia criterion provides straightforward guidance to identify the number of clusters from the mathematical perspective, our analysis showed that such a recommendation is not necessarily optimal from the application standpoint; repeating the analysis with a few different values of K and comparing and contrasting the results will lead to well-informed conclusions. Second, confidence on the risk indices rendered by our model directly relies on the number of observations for each cluster, independent of the others. For example, the inference of *r* = 0.0% for the first and second regions of the dose-volume space in Figure 4b is made based on 9 and 8 observations, respectively.

While this estimate is acceptable for these low-dose regions, in general, such small samples are highly sensitive to noise. Finally, ORN has low prevalence, which makes the classes of ORN and non-ORN patients unbalanced. This poses a challenge to statistical inference, in general, both supervised- and unsupervised-learning methods.

Visualization tools play a key role in clinical interpretability and reliability of mathematical models. In our analysis, visualization of the identified clusters by the K-means clustering method uncovered the distinction between the sixth and first five clusters and led to an improved partitioning of the dose-volume space. Visualization of the intermediate results may also provide additional insight for radiation planning. For example, the sixth (black) cluster may be named the cluster of *post-operative radiation plans*; more than 77% of the patients in this cluster have received post-operative radiation therapy while this percentage for the other clusters remains below 20%. The special shape of this cluster is easily detectable to oncologists, which immediately reveals the relatively high prevalence of ORN among this group of patients.

Availability of a visualization dashboard to illustrate every step of the process (Figures 1 to 3)— rather than just the final result (Figure 4)—can greatly improve clinical reliability of the proposed analysis for risk evaluation of ORN (with different data sets) and other complications.

## 5. Conclusion

This paper presents the results of an unsupervised-learning analysis for mandibular ORN risk evaluation on retrospective data of a large cohort of HNC patients treated by radiation therapy at the University of Texas MD Anderson Cancer Center between 2005 and 2015. The presented analysis provides a visual tool for ORN risk assessment and a spectrum of dose constraints on the mandible for radiation planning with respect to this complication. The unsupervised-learning approach is extendable to other radiation therapy side effects to improve the current state of NTCP modeling.

## Data Availability

All research data are publicly available at https://doi.org/10.6084/m9.figshare.13568207

## Appendix A: Effect of Data Inseparability on Classification Accuracy

**Figure A.1:**
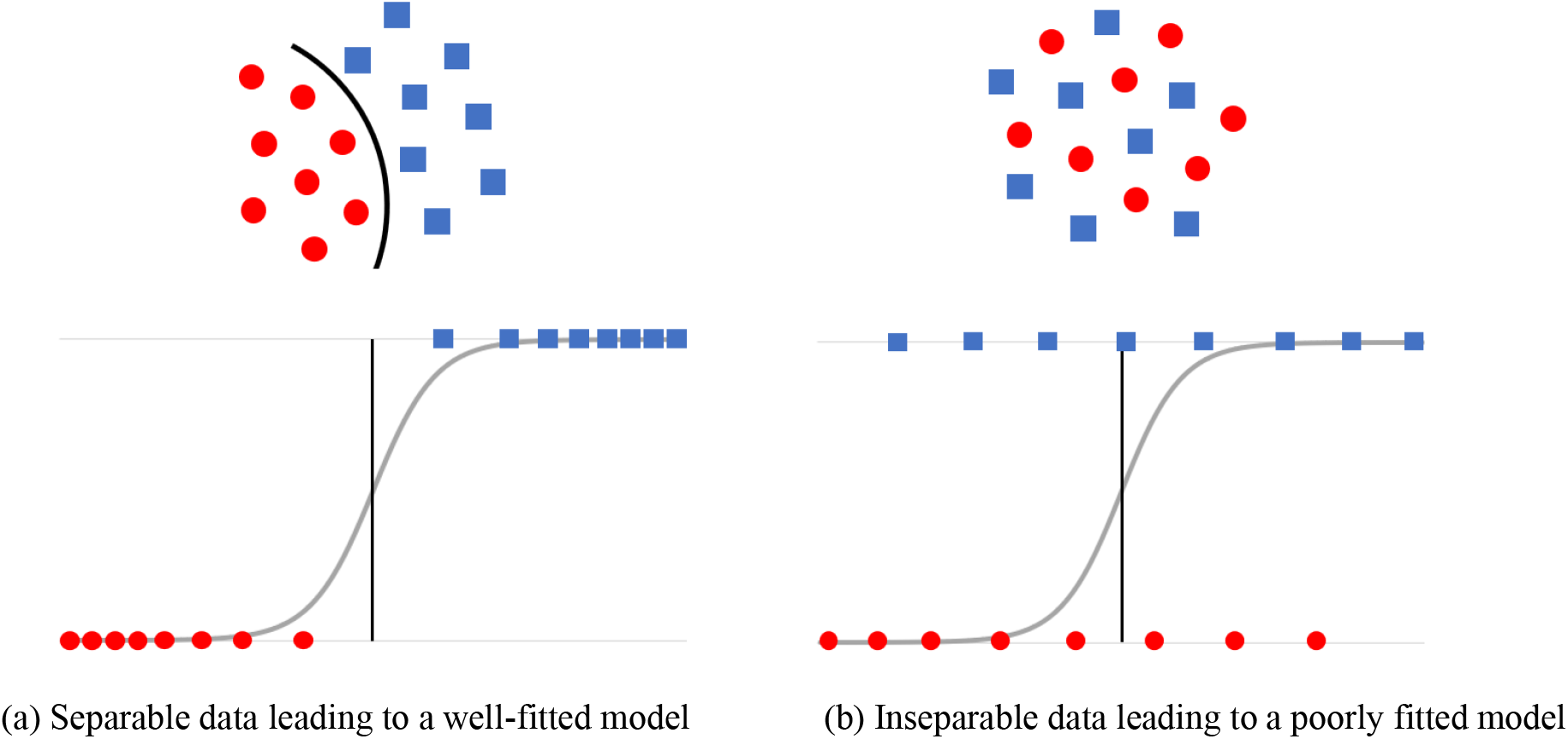
Data inseparability and its effect on logistic regression performance

## Appendix B: ORN Data Inseparability by DVH Features

The following figures show that the two classes of ORN-negative and ORN-positive patients in our data set are inseparable by D30% and pre-radiation dental extraction (PDE) status. Given only two variables, this could be shown on a single plot. However, for better visualization and providing more insight on the structure of the data, we illustrate the data inseparability in smaller groups; that is, for each pair of DVH cluster and PDE status separately. Evidently, if the whole data set is separable, it remains separable when divided in smaller groups. Also, for each patient, we have plotted D30% against the standard deviation (SD) of the delivered dose, for visualization purposes only, so all data points are not located on a horizontal line.

**Figure B.1:**
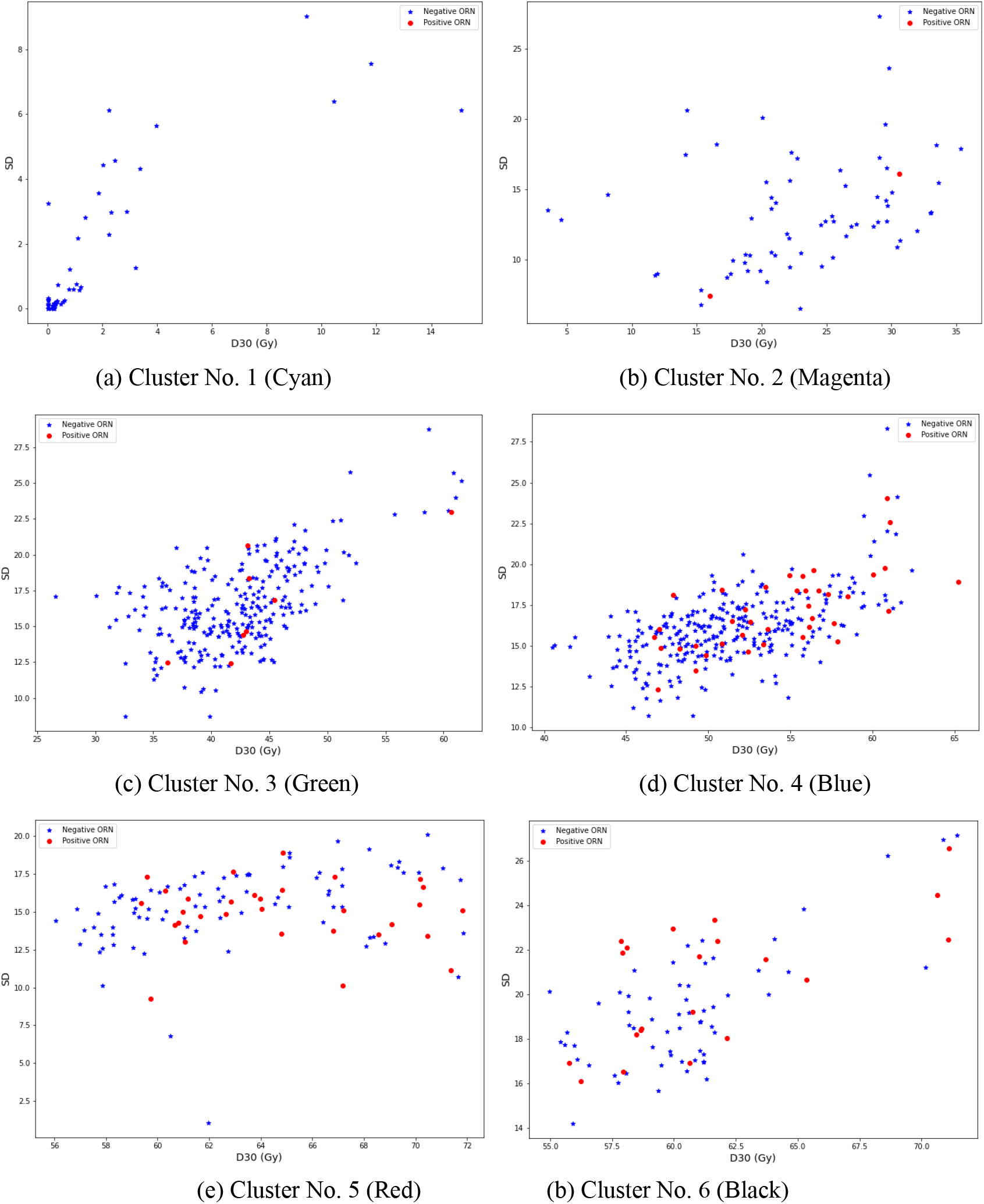
Data inseparability within the clusters for PDE = 0

**Figure B.2:**
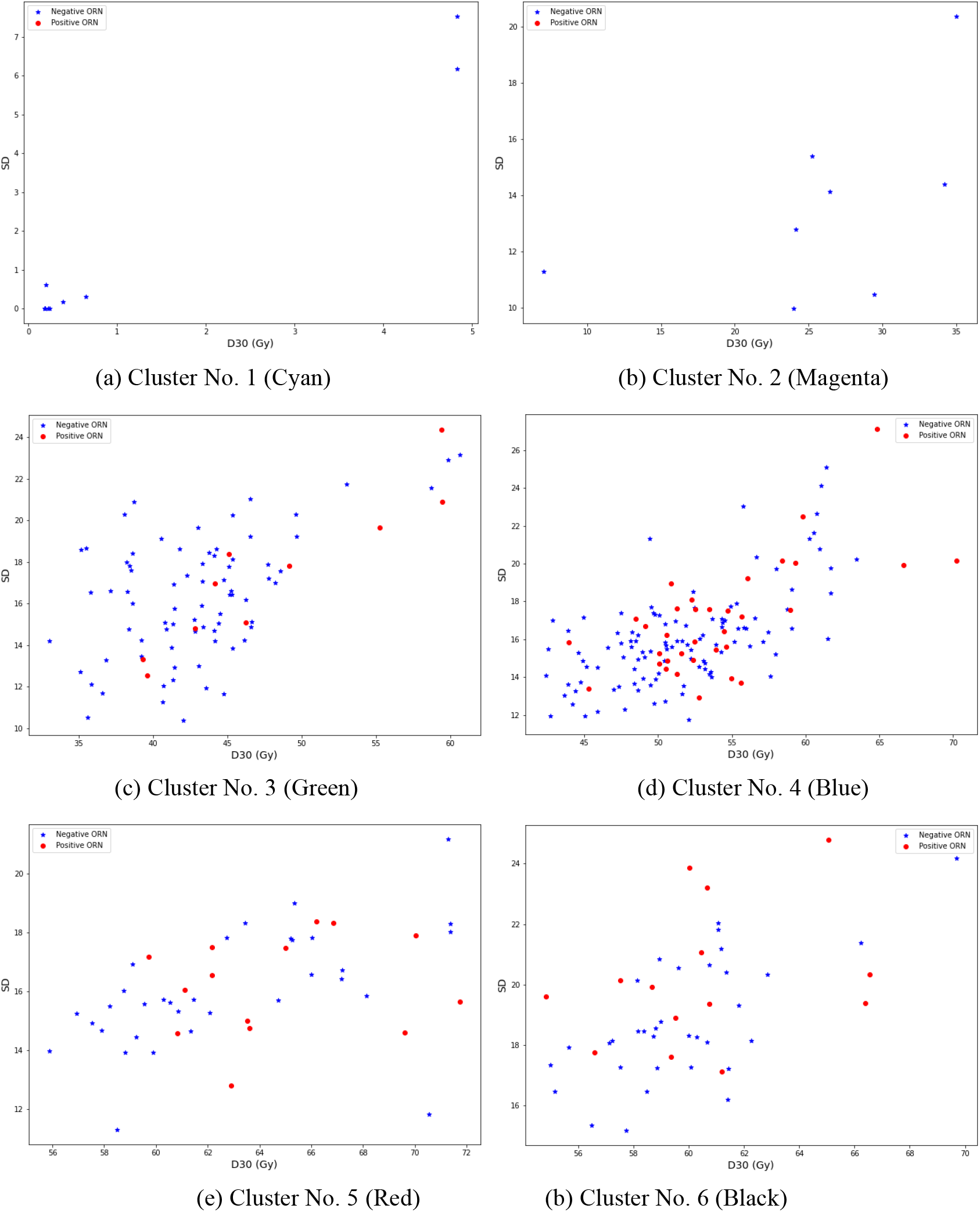
Data inseparability within the clusters for PDE = 1

Figures B.1 and B.2 show that, in all D30%-PDE categories, the class of ORN-negative patients is dominant, and the two classes are highly mixed (inseparable). As we have used SD (y axis) for better visualization, the data inseparability by D30% (for each PDE status) is implied by nonexistence of vertical separators of “Negative ORN” and “Positive ORN” points. However, the fact that these classes cannot be separated by other lines/curves implies that including SD as a feature is not expected to improve the performance of a logistic regression model. These figures also reveal that introducing “DVH cluster” as an independent feature to a logistic regression model is not likely to significantly improve its performance, due to inseparability of data in all D30%-cluster-PDE categories. Finally, the existence of high (linear) correlations among Dx% features implies that replacing D30% with any other DVH feature (including the mean dose) will not improve the inseparability.

## Appendix C: Logistic Regression vs. Naive Bayes

**Figure C.1:**
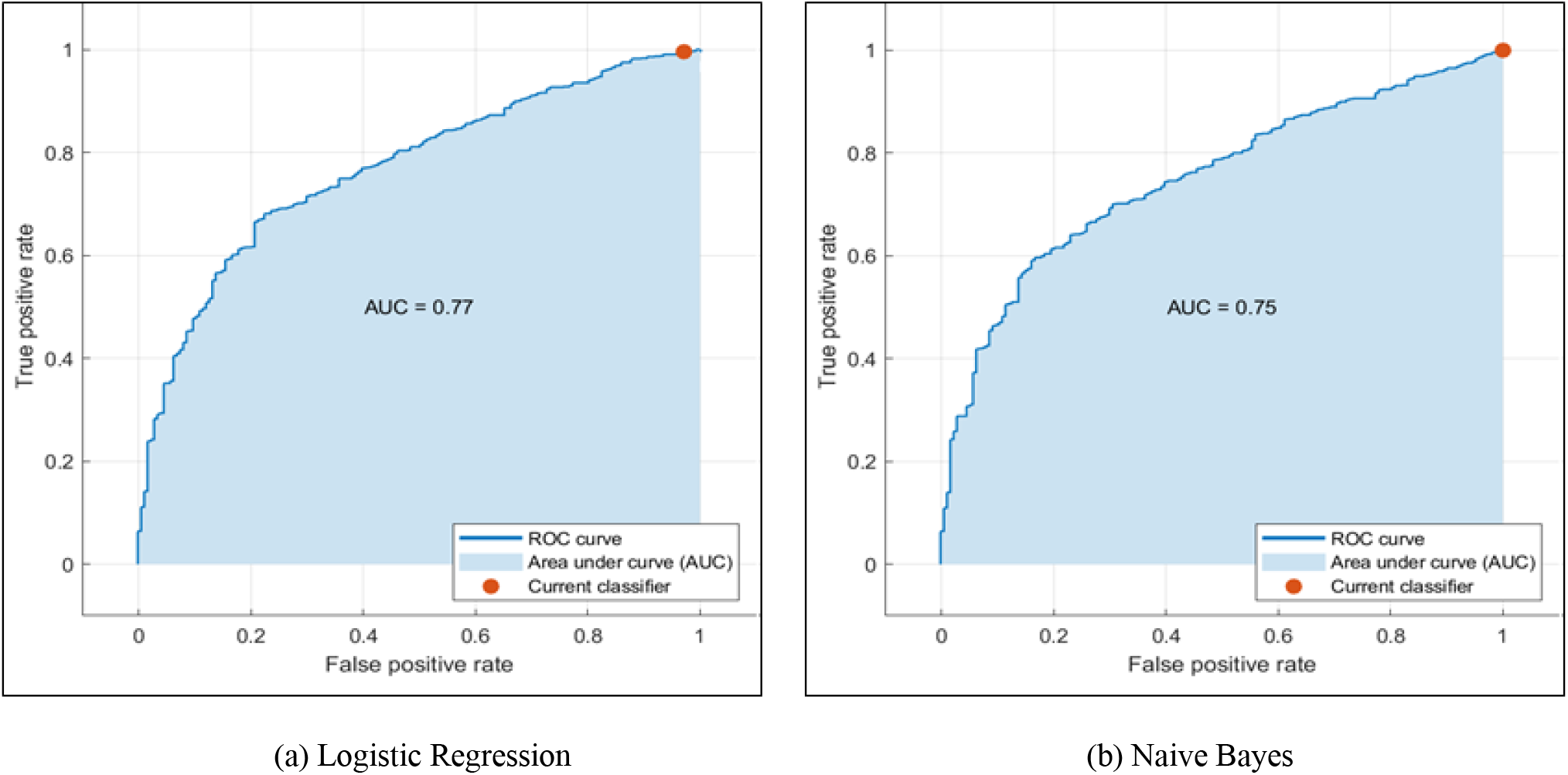
Comparison of the performances of logistic regression and naive Bayes on the data (The features include D30% and PDE status for both models.)

